# Benign Bone Lesions: Do They Warrant Follow-Up?

**DOI:** 10.1101/2023.06.20.23291609

**Authors:** Andrew Brook, Harrison Volaski, Emily Kleinbart, Jichuan Wang, Swapnil Singh, Rui Yang, Bang Hoang, Waleed Al-Hardan, Ranxin Zhang, Beverly Thornhill, David S. Geller

**Author notes:** Corresponding author: David S. Geller, MD, Department of Orthopaedic Surgery, Montefiore Health System 3400 Bainbridge Avenue, Bronx, NY 10467, USA.

## Abstract

**Background:** There are no established surveillance guidelines for benign bone lesions, particularly for those that do not merit surgery. It is unclear how long or how often patients should be followed, what type of radiographic studies should be obtained, and how frequency repeat imaging should be performed. Given that follow-up incurs cost, time, and resources, it is essential to better understand the probability of lesion progression and the necessity, or lack thereof, for clinical and radiographic observation.

**Methods:** A retrospective review was conducted between 2015 and 2020 of patients of all ages, races, and sexes diagnosed with a benign bone lesion after radiographic imaging.

Patients diagnosed with benign bone lesions outside of the study period or not managed by an orthopedic surgeon were excluded. Outcomes included presenting symptoms, the date of visits to an orthopedic surgeon, imaging, the appearance or type of lesion, and lesion location. Patients were divided into two groups, those who were observed (Group 1) and those who underwent surgery during the duration of the study (Group 2). Both groups were subdivided into patients who were asymptomatic (Group 1a and Group 2a) or symptomatic at presentation (Group 1b and Group 2b). Descriptive statistics were used to analyze the data extracted.

**Results:** Of the 638 patients included, 10 patients (1.6%) demonstrated a change in either lesion size or morphology, 9 of which were pediatric patients. Patients in Group 1a were followed, on average, for 207.0 days and returned to the office 1.3 times after their initial visit. Patients in Group 1b were followed, on average, for 130.0 days and returned to the office 1.4 times after their initial visit. Patients in Group 2a were followed, on average, for 191.8 days and returned to the office 1.4 times after their initial visit. Patients in Group 2b were followed, on average, for 102.0 days and returned to the office 1.2 times after their initial visit. The most common imaging study obtained were plain radiographs. Patients in Group 1a received repeat imaging studies, on average, every 100.7 days while patients in Group 1b received repeat imaging studies, on average, every 69.3 days. Patients in Group 2a received repeat imaging studies, on average, every 90.3 days while patients in Group 2b received repeat imaging studies, on average, every 47.3 days.

**Conclusions:** Benign bone lesions are common incidental findings, and most require no surgical intervention. There are currently no guidelines for how long and how frequently patients should be followed, either clinically or radiographically. This study demonstrates that progression is an extremely uncommon event. Moreover, when progression does occur, it is often accompanied by clinical symptomatology. Limiting clinical and radiologic follow-up to symptomatic individuals would save most patients from incurring costs related to unnecessary clinical visits and repeat imaging studies and reduce their overall lifetime exposure to radiation. In an increasingly resource-challenged environment, routine sequential follow-up may be hard to justify. Reassuring patients and parents that access is available, if and when needed, may be helpful in managing concern while limiting cost and exposure.

## Introduction

### Background

Benign bone lesions can develop in any part of the human skeleton and can develop at any age, though they typically predominate in children and adolescents [28]. Most are found incidentally and do not require intervention. Nevertheless, patients are often referred to an orthopedic oncologist for evaluation and management [26]. It has been suggested that as many as 57% of new consultations in an orthopedic oncology practice concern benign bone lesions, and their overall prevalence is thought to approach 19% within an asymptomatic population, though this is likely an underestimation [9]. Although intervention is sometimes warranted [24], many benign bone lesions are treated nonoperatively with observation [23].

In fact, many of these have been termed “leave me alone” lesions, and some have even been recognized as being self-limiting [25]. Regardless, these lesions can be anxiety-provoking for patients and their family members, who frequently require explanation and reassurance.

Providers may be inclined to repeatedly evaluate the patient or image the lesion to provide maximal reassurance. Moreover, providers may practice defensively, ordering multiple studies to objectively confirm the lesion exhibits stable, non-aggressive features [24]. Finally, there is a paucity of literature that clearly outlines or quantifies the likelihood of benign bone lesion growth or progression [9], which probably contributes to varying practice styles or preferences.

Currently, there are no universal guidelines for how often or in what manner a patient should be observed. For example, it remains unclear how often patients should be seen for a clinical evaluation, how frequently they should undergo repeat imaging studies, what type of imaging studies should be obtained, and how long follow-up should last. This can lead to multiple follow-up visits and repeated imaging of uncertain value, potentially resulting in added cost, time expenditure, anxiety, and radiation exposure [26]. Follow-up often requires time away from either school or work and demands both provider time and medical resources [11], which might otherwise be better directed. Although a single lesion incurs minimal cost, the cumulative impact across a population or a health care system can quickly balloon, underscoring the matter’s relevance and the need to better understand current practice.

Toward this end, we sought to assess the natural course of benign bone lesions and their management within the context of a single academic medical center. We asked the following questions: 1) How often do incidentally found bone lesions or bone lesions not requiring surgery at presentation progress over time? 2) What is the length of follow-up for these patients and the frequency of follow-up visits within our institution? and 3) What imaging studies were obtained and how frequently were they repeated?

## Methods

### Study design and setting

A single-institution retrospective review was conducted at a tertiary care urban healthcare system and included patients diagnosed with a benign bone lesion between January 1, 2015, and December 31, 2020.

### Participants/study subjects

Patients were identified using the Observational Health Data Sciences and Informatics (OHDSI) ATLAS web application, which yielded a list of 2546 patient MRNs. Each patient chart was individually reviewed by one of three investigators to assess whether inclusion criteria were met. Patients met inclusion criteria if diagnosed with a new benign bone lesion on radiographic imaging within the aforementioned time frame and if they also were evaluated by an orthopedic surgeon. All sexes, ages, and races were included. Patients were excluded if they were diagnosed with an entity other than a benign bone lesion, if they were diagnosed with a benign bone lesion prior to the study period, if they were indicated for surgery at their first visit, or if they returned to the office to be reevaluated after not being seen for over two years. Data was managed using a HIPAA-compliant password-protected encrypted database. Institutional review board approval was obtained (IRB #: 2021-12664).

### Variables, outcome measures, data sources, and bias

Data points included basic demographics and information obtained at the initial and follow-up visits, including the date of the initial complaint, the reason for seeking medical attention, the date of the first orthopedics evaluation, if the lesion was noted incidentally, symptomatology, ultimate diagnosis, and the anatomic location, size, and morphology of the lesion. Information regarding imaging studies included the date of the imaging study, the type of imaging, how many radiographic views were obtained, if sedation was used, the imaging findings, and if additional studies were ordered. If a biopsy or a surgery was performed, the date, type, and results were recorded. If the patient underwent surgery, we noted if it was an index procedure or a re-operation. The time between visits and between imaging studies was noted as well.

Definitive outcomes were classified as ‘Resolution of Lesion,’ ‘Active Surveillance,’ ‘Discharged from Follow-Up,’ ‘Surgery,’ ‘Radiofrequency Ablation,’ and ‘Did not Seek Further Follow-Up.’ ‘Resolution of Lesion’ was defined by radiographic resolution of the lesion noted in the patient’s chart and imaging report. ‘Active Surveillance’ was defined as patients actively seeking follow-up and currently have upcoming appointments scheduled. ‘Discharged from Follow-Up’ was defined as patients whose charts indicated that follow-up was no longer recommended. ‘Surgery’ was defined as patients who underwent an excision, biopsy with subsequent curettage & packing, curettage & packing, or open reduction and internal fixation (ORIF). ‘Radiofrequency ablation’ was defined as patients undergoing radiofrequency ablation. ‘Did not Seek Further Follow-Up’ was defined as patients who were instructed to return but never did.

If the patient was not indicated for interventions at presentation, they were considered as “under observation.” Length of follow-up was defined as the time between the patient’s first visit with an orthopedic surgeon and their last follow-up visit. A change in size or morphology of the lesion was determined by radiographic reports indicating that the lesion had grown or otherwise evolved.

Given the incidental and asymptomatic nature of these lesions, it is not uncommon for patients to forget their appointments or choose to not follow-up. An effort was made to collect longer follow-up data via a telephone survey for non-operative patients. Patients that had been lost to follow up were called, and long-term follow-up data was collected on 79 of those 377 patients. Patients or legal guardians were consented over the phone. A full cohort data analysis and a sub-cohort analysis of pediatric patients (age 0 to18) and adult patients (age 19 to 86) were conducted. The full list of questions is included in the supplemental section.

### Accounting for all patients / study subjects

Of the 2546 patients initially identified, 1908 patients were excluded because they were given a diagnosis other than a benign bone lesion (1178 patients), they were diagnosed prior to January 1, 2015 (271 patients), their charts did not provide complete datasets (91 patients), or they were never seen by an orthopedic surgeon (81 patients), leaving 925 patients who met inclusion criteria. All cases were initially reviewed, and analysis was focused on a subset of patients with the 5 most common histological subtypes which comprised a cohort of 638 patients. Patients were divided into a pediatric group of patients aged 0 to 18 (Group 1, n=278), and an adult group of patients aged 19 to 86 (Group 2, n=360). Group 1 and Group 2 were subdivided further into patients who were asymptomatic at presentation, Group 1a (n = 89) and Group 2a (n = 219), respectively, and those who were symptomatic at presentation, Group 1b (n = 189) and Group 2b (n = 141), respectively (Figure 3).

### Demographics, description of study population

This cohort was managed either initially or entirely by observation. It demonstrated a slight female predominance (52.0%), and the average age was 31.9 ± 21.5 years (Table 1). Ages ranged from less than 1 to 86, with most patients diagnosed in the second decade of life (Figure 1).

**Figure 1.**
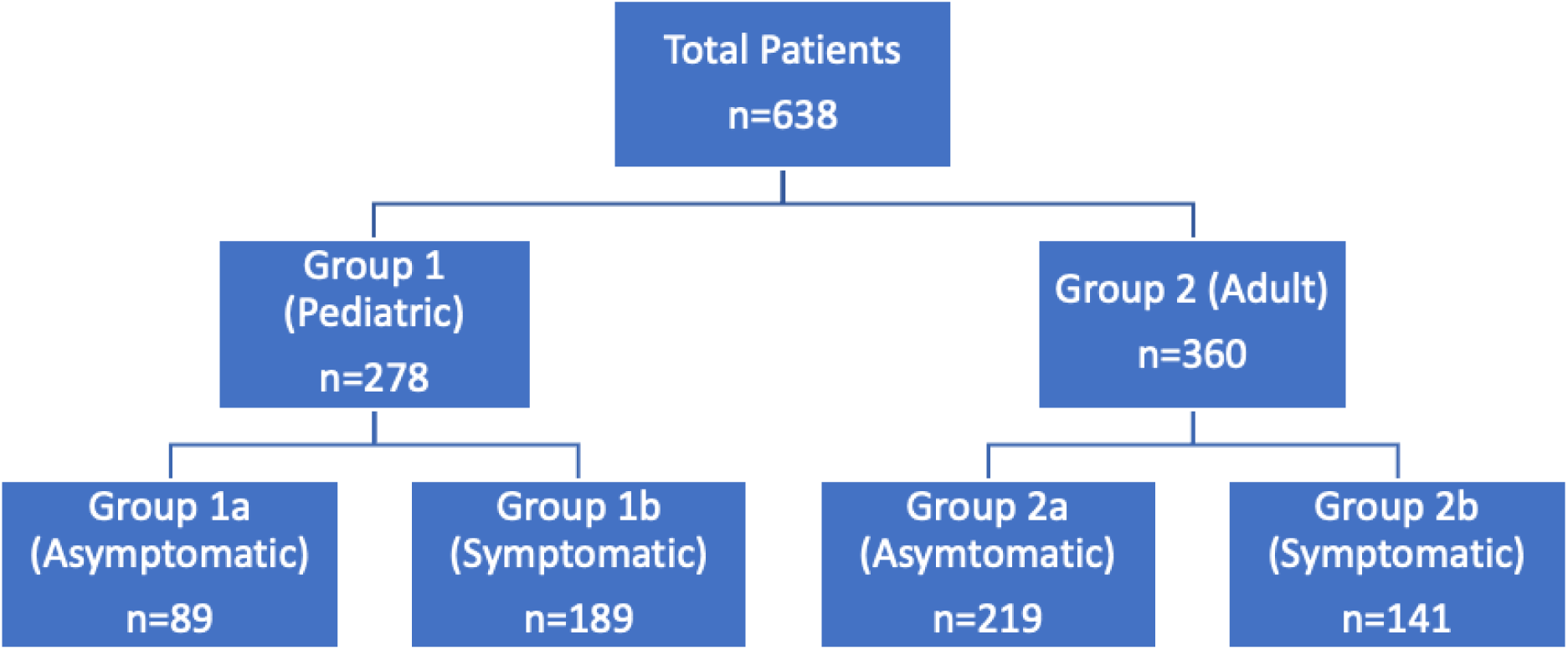
Cohort overview. Adult and pediatric patients were analyzed separately and were subsequently split into two groups based on symptomatology at presentation.

**Table 1.** Patient demographics.

|  |  |  |
| --- | --- | --- |
| Gender | 52.1% Female | 47.9% Male |
| Age | 31.9 ± 21.5 years |  |
| Race | 10.5% White |  |
|  | 16.6% Black |  |
|  | 36.1% Hispanic |  |
|  | 1.0% Asian |  |
|  | 11.4% Other |  |
|  | 25.2% Declined |  |

### Statistical analysis, study size

The incidence of benign bone lesion progression at our institution was calculated for the entire cohort of patients included in this report. Additionally, a sub-analysis was performed on the telephone survey cohort. Analysis groups were not compared, so statistical tests were not employed. Descriptive statistics were utilized to quantify and describe patient characteristics and trends in the results.

## Results

Thirty different types of benign bone lesions were identified in the patients that met inclusion criteria, and the 5 most common diagnoses were osteochondroma (23.4%), enchondroma (23.0%), non-ossifying fibroma (NOF) (13.2%), unicameral bone cyst (5.5%), and monostotic fibrous dysplasia (4.1%) (Figure 2). The most involved anatomic locations were the femur, followed by the tibia (Table 2).

**Figure 2.**
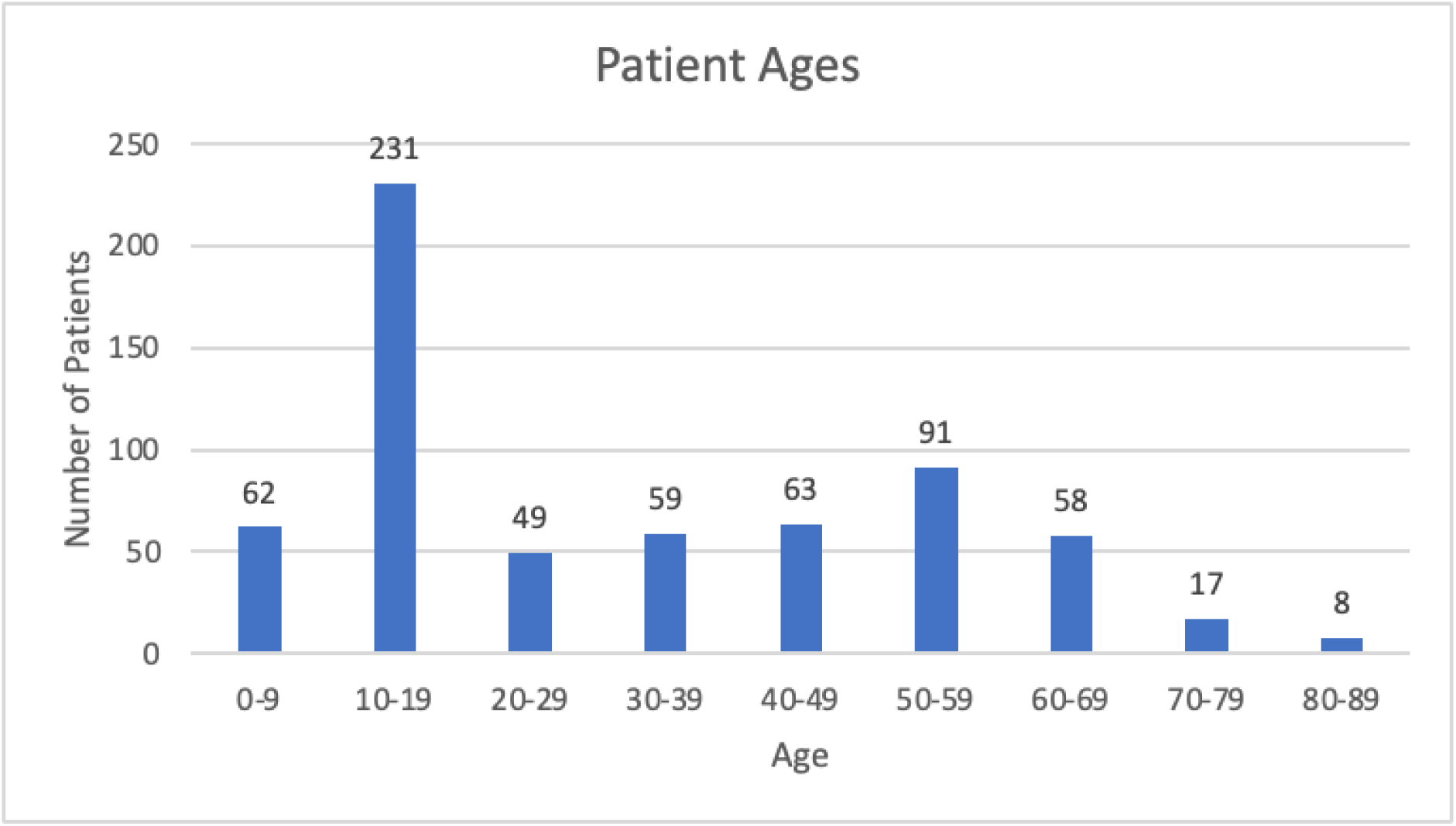
Patient ages. The average patient age was 31.9 ± 21.5 years. Ages ranged from less than 1-year-old to 86-years-old. The majority of patients were diagnosed in the second decade of life.

**Figure 3.**
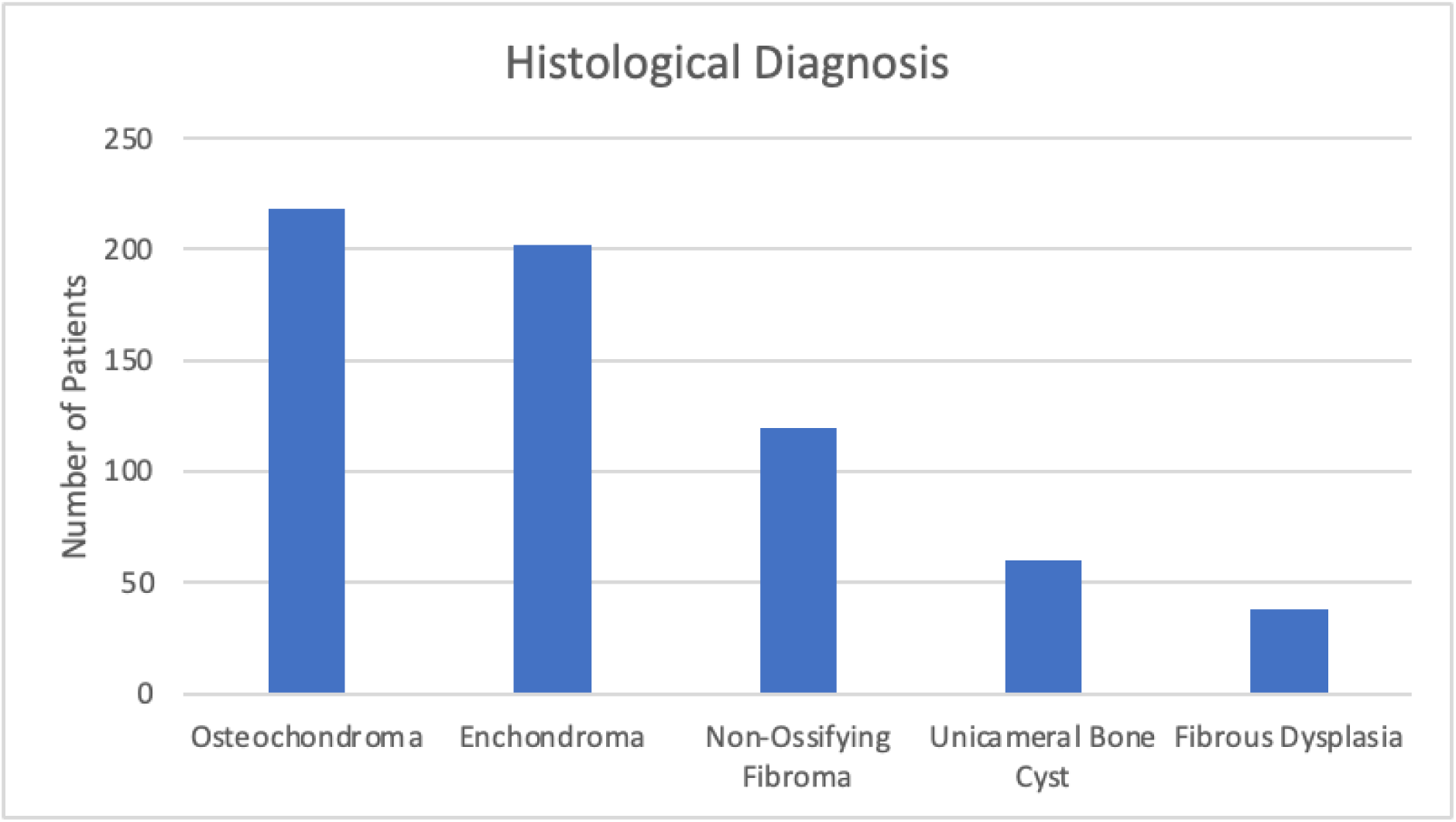
Benign bone lesions diagnosed. Osteochondromas and enchondromas made up 65.8% of the entire cohort.

**Table 2.**

| Location | Percent of Patients |
| --- | --- |
| Femur | 38.93 |
| Tibia | 25.04 |
| Humerus | 15.73 |
| Hand | 7.18 |
| Fibula | 3.97 |
| Scapula | 2.14 |
| Foot | 1.98 |
| Pelvis | 1.53 |
| Radius | 1.53 |
| Ulna | 0.61 |
| Ribs | 0.61 |
| Spine | 0.31 |
| Clavicle | 0.31 |
| Manubrium | 0.15 |

### How often do incidentally found bone lesions or bone lesions not requiring surgery at presentation progress over time?

Within the entire cohort of 638 patients, only 10 (1.6%) demonstrated a change in either lesion size or morphology, 9 of which were pediatric patients. One adult patient exhibited findings most compatible with fibrous dysplasia, and 7 of the 9 pediatric patients demonstrated osteochondromas (Table 3). Seven of the 10 reported symptoms at presentation, and 7 of the 10 were ultimately managed operatively. Of the seven patients treated surgically, three reported new or worsening pain due to the lesion, two requested surgical management owing to anxiety or cosmesis, 1 sustained a pathological fracture, and 1 exhibited persistent lesion growth near the proximal humerus epiphysis prompting the recommendation to manage operatively. Of the remaining three patients demonstrating lesion progression, one was lost to follow-up, and two remain asymptomatic and are under ongoing observation. The average length of time between presentation and noted lesion change was 400 days, or just over 13 months. Lesion growth was diagnosed on plain radiographs for 9 patients, and on MRI for one.

**Table 3.** Patients with increase in lesion size.

| Diagnosis | Age Range | Sex | Bone | Symptoms at presentation | Changes in symptoms/<br>Reason for surgery | When change was noticed |  | Imaging modality | Outcome |
| --- | --- | --- | --- | --- | --- | --- | --- | --- | --- |
|  |  |  |  |  |  | Follow up visit | Days |  |  |
| Osteochondroma | 11-15 | M | Femur | None | None | 1 | 504 | XR | Active Surveillance |
| Osteochondroma | 11-15 | M | Femur | Mass | None | 1 | 231 | XR | Active Surveillance |
| Osteochondroma | 11- 15 | F | Femur | Mass | None | 1 | 315 | XR | Lost to Follow Up |
| Osteochondroma | 11-15 | M | Femur | None | Worsening pain | 1 | 511 | XR | Surgery |
| Osteochondroma | 11-15 | F | Fibula | Pain | Continual pain with visible/palpable growth | 2 | 693 | XR | Surgery |
| Osteochondroma | 6-10 | F | Tibia | Pain and mass | Patient's guardian requested removal | 1 | 49 | XR | Surgery |
| Osteochondroma | 11-15 | M | Tibia | Pain and Mass | Patient's guardian requested removal | 2 | 322 | XR | Surgery |
| Unicameral Bone Cyst | 6-10 | M | Humerus | Fracture | Lesion growth near growth plate | 2 | 280 | XR | Surgery |
| NOF | 16-20 | F | Tibia | None | Unrelated fracture near lesion requiring surgical repair | 3 | 908 | XR | Surgery |
| Fibrous Dysplasia | 41-45 | F | Tibia | Pain | Worsening Pain | 3 | 182 | MRI without contrast | Surgery |

In the extended follow-up survey, 4 patients (5.1%) subjectively reported feeling their lesion had grown, and 2 (2.5%) felt that it had healed. Of the 4 that felt their lesion had grown, 2 reported undergoing imaging studies at an outside institution that showed their lesions were stable. Twenty-one patients in total reported obtaining additional imaging at outside institutions. One NOF was found to have increased in size on imaging obtained for unrelated acute joint pain (1.3%) (Table 4).

**Table 4.** Telephone survey.

|  | **Pediatric Cohort (n=29)** | **Adult Cohort (n=50)** |
| --- | --- | --- |
| **Demographics** |  |  |
| Males (%) | 15 (51.7) | 15 (30) |
| Average age | 13.3 ±18.0 | 42.8±17.8 |
| Average number of years since last follow-up (range) | 5 (2-8) | 5 (2-8) |
| **Diagnosis** |  |  |
| Osteochondroma (%) | 13 (44.8) | 11 (22.0) |
| Nonossifying Fibroma (%) | 12 (41.4) | 7 (14.0) |
| Unicameral bone cyst (%) | 2 (6.9) | 0 (0.0) |
| Enchondroma (%) | 1 (3.4) | 28 (56.0) |
| Fibrous Dysplasia (%) | 1 (3.4) | 4 (8.0) |
| **Patient-Reported Responses** |  |  |
| New symptoms (%) | 7 (24.1) | 16 (32.0) |
| Surgery (%) | 1 (3.4) | 0 (0.0) |
| Sought care from another institution (%) | 7 (24.1) | 7 (14.0) |
| Subjective growth (%) | 1 (3.4) | 3 (6.0) |
| Subjective healing (%) | 1 (3.4) | 1 (2.0) |
| Repeat imaging obtained (%) | 9 (31.0) | 12 (24.0) |
| Imaging for lesion (%) | 6 (20.7) | 4 (8.0) |
| Unrelated imaging (%) | 3 (10.3) | 8 (16.0) |
| Radiographic evidence of growth (%) | 1 (3.5) | 0 (0.0) |
| Radiographic evidence of healing (%) | 3 (10.3) | 0 (0.0) |
| Additional lesion found on imaging (%) | 0 (0.0) | 2 (4.0) |
| New cancer diagnosis (%) | 0 (0.0) | 0 (0.0) |
| Anxiety due to lesion (%) | 5 (17.2) | 15 (30.0) |

When questioned about symptoms in the telephone survey, 21 patients reported pain (26.6%), 3 reported swelling at the lesion site (3.8%), and no patients sustained a pathological fracture since their last follow-up visit. Of the patients that reported having pain, 14 had chronic pain that persisted since their original diagnosis, and 7 had new onset pain since their last follow-up visit. However, 13 of the 21 patients that reported pain exhibited medical information that strongly suggested an alternative etiology, with 7 having sustained unrelated injuries to the area and 6 with chronic osteoarthritis, leaving only 8 patients (10.1%) with pain that could not otherwise be attributed to non-lesion related causes.

### What is the length of follow-up for patients diagnosed with a benign bone lesion and the frequency of follow-up visits within our institution?

Of the patients in Group 1, those in Group 1a returned to the office, on average, 1.3 times after their initial visit, compared to an average of 1.4 times for patients in Group 1b (Figure 4). While many patients did not return for follow-up at all, approximately 68% of both asymptomatic patients and symptomatic patients returned for at least one follow-up visit. The average length of follow-up was 207.0 days and 130.0 days for patients in Group 1a and Group 1b, respectively. The average length of time between follow-up visits was 88.0 days for patients in Group 1a and 68.8 days for those in Group 1b.

**Figure 4.**
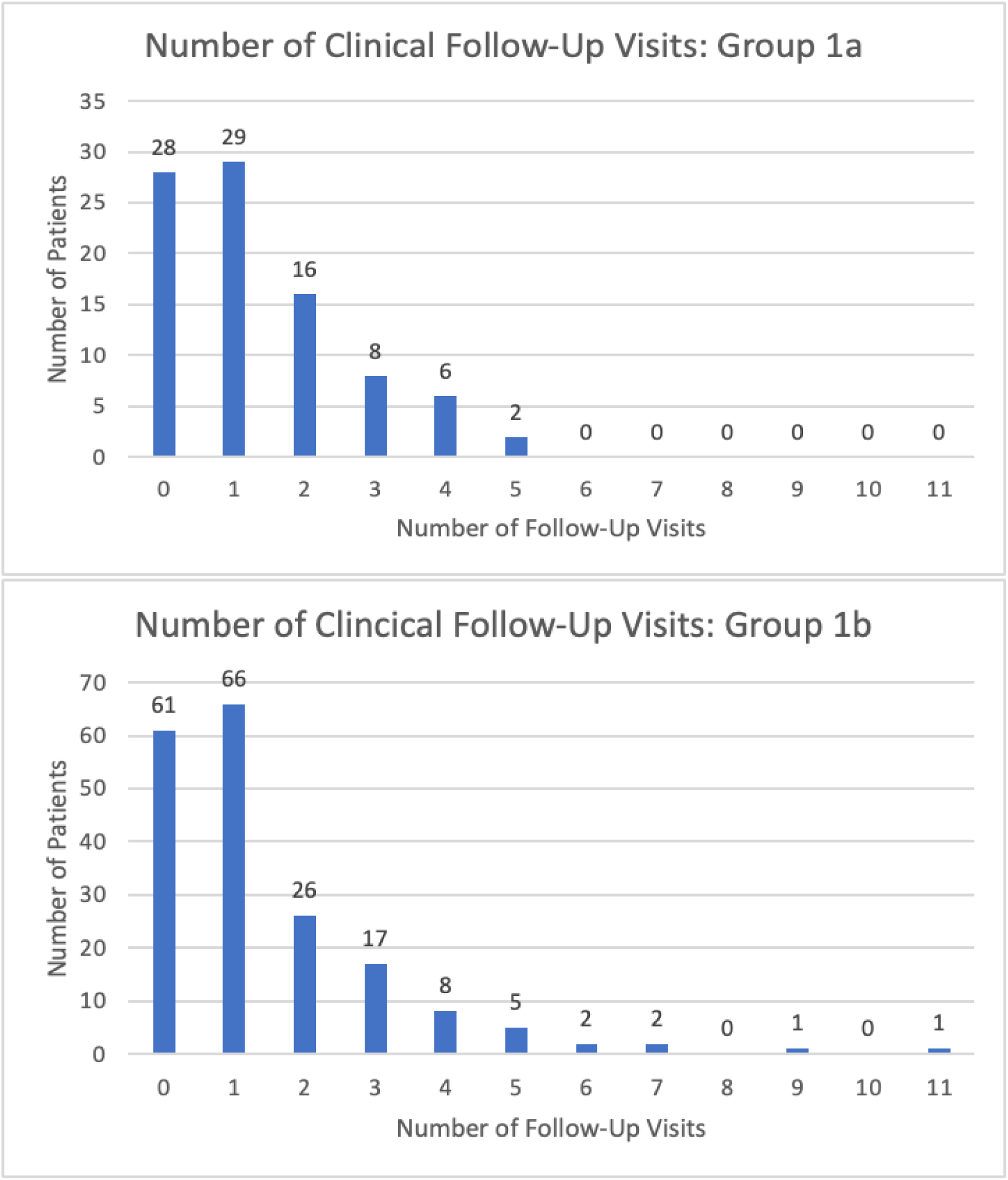
Number of follow-ups for patients in Group 1. A) Patients in Group 1a came for an average of 1.3 clinical follow-up visits. The average length of time between follow-up visits was 88.0 days, and the average total length of follow-up was 207.0 days. B) Patients in Group 1b came for an average of 1.4 clinical follow-up visits. The average length of time between follow-up visits was 68.8 days, and the average total length of follow-up was 130.0 days.

Within Group 2, those in Group 2a returned to the office, on average, 1.4 times after their initial visit, while those in Group 2b returned on average, 1.2 times (Figure 5). The average length of follow-up was 191.8 days for patients in Group 2a and 102.0 days for patients in Group 2b. The average length of time between follow-up visits was 85.0 days for patients in Group 2a and 50.1 days for patients in Group 2b.

**Figure 5.**
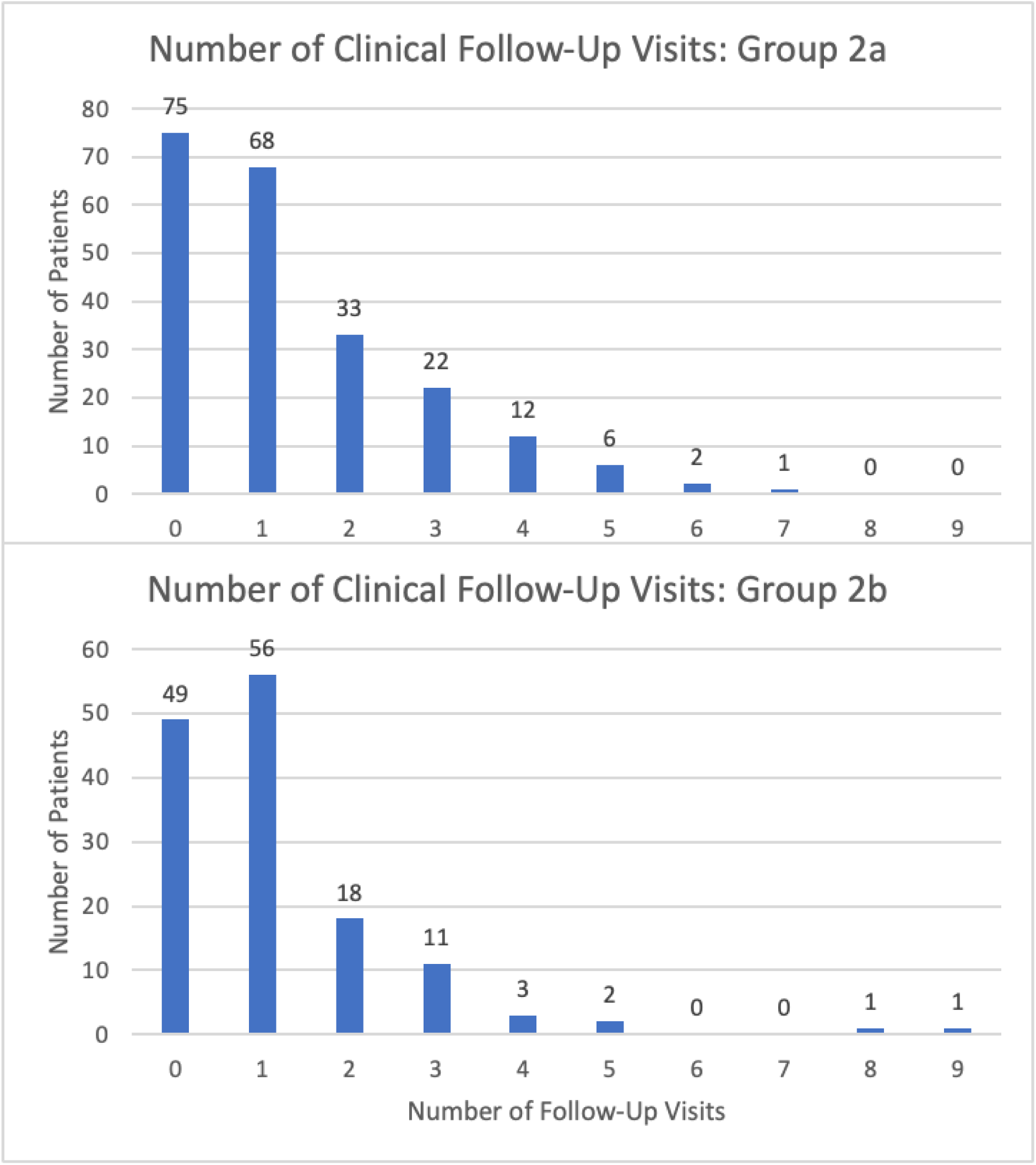
Number of follow-up visits for patients in Group 2. A) For patients in Group 2a, the average number of follow-up visits after the patient’s first visit with an orthopedic surgeon was 1.4 visits, the average length of time between follow-up visits was 85.0 days, and the average total length of follow-up was 191.8 days. B) For patients in Group 2b, the average number of follow-up visits was 1.2 visits, the average length of time between visits was 50.1 days, and the average length of follow-up was 102.0 days.

Of the patients that were symptomatic at presentation, 97 (51%) of the patients in Group 1b and 72 (51%) of the patients in Group 2b required surgery (Figure 6). The most common symptom patients in both groups experienced was pain (Figure 7). Of the patients that were asymptomatic at presentation, 10 (11%) of patients in Group 1a and 12 (5%) of patients in Group 2a eventually required surgery. The 10 originally asymptomatic pediatric patients that required surgery all developed symptoms over the course of their observation, and the most common indication for surgery in this group was pain (60%). Of the 12 originally asymptomatic adult patients that required surgery, 7 of them developed symptoms during the course of their observation, and 5 remained asymptomatic but required surgery either for diagnostic purposes, impending fracture risk, or unrelated procedures that necessitated the removal of the lesion (eg, a lesion impacting planned hip replacement).

**Figure 6.**
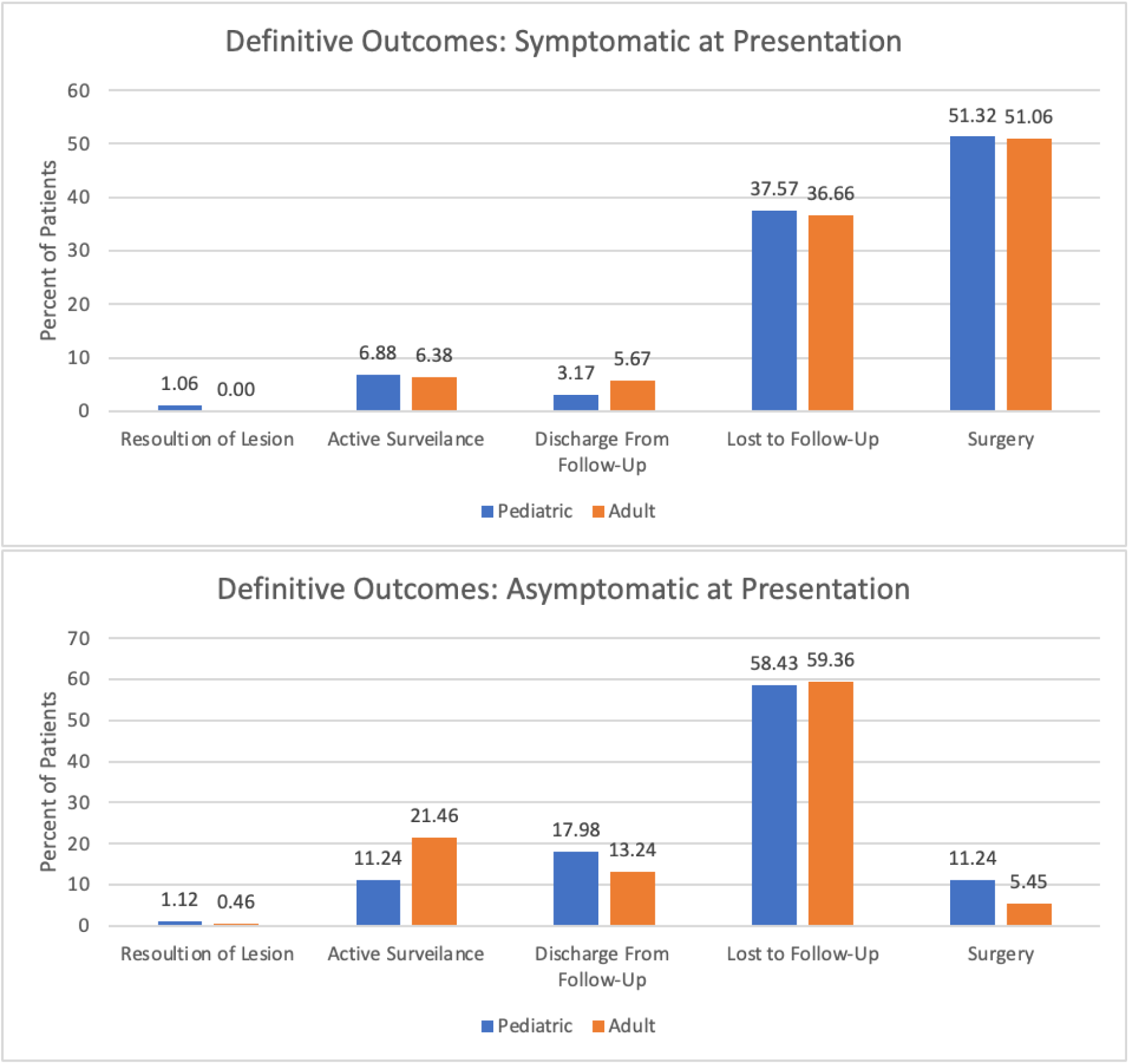
Definitive outcomes. Most patients did not return for follow-up. ‘Resolution of Lesion’ was defined by radiographic resolution of the lesion noted in the patient’s chart and imaging report. ‘Active Surveillance’ was defined as patients who were actively seeking follow-up and currently have upcoming appointments scheduled. ‘Discharged from Follow-Up’ was defined as patients whose charts indicated that follow-up was no longer recommended.

**Figure 7.**
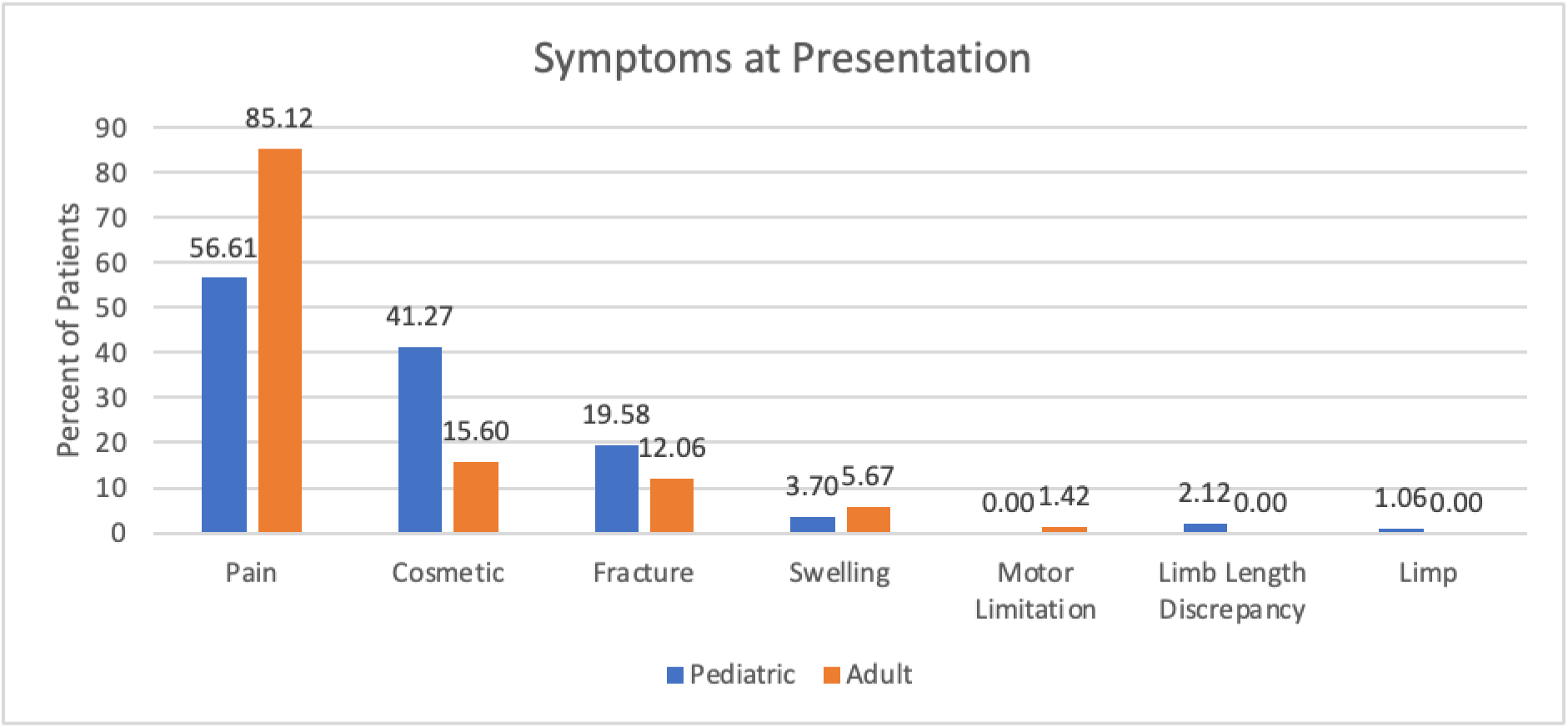
Symptoms at presentation. This chart displays the symptoms patients in Groups 1b and 2b complained of at their first visit with an orthopedic surgeon. The most symptom was pain. “Cosmetic” was defined as a visible or palpable mass that was bothering the patient cosmetically or causing them anxiety about the lesion.

### What imaging studies are obtained, and how frequently are repeat imaging studies obtained?

Approximately 60.1% of patients in Group 1 had at least one follow-up imaging study. Patients in Group 1a had 1, 2, or more than two studies in 29.2%, 13.5%, and 15.7% of cases, respectively. Patients in Group 1b comparatively underwent 1, 2, or more than two additional studies in 34.9%, 12.7%, and 13.2%, respectively (Figure 8). The average length of time between studies was 100.7 days for patients in Group 1a and 69.3 days for patients in Group 1b. The average length of time between repeat imaging studies was 90.3 days for patients in Group 2a and 47.3 days for patients in Group 2b. Within Group 2, 58.9% of patients had at least one follow-up imaging study taken. Patients in Group 2a had 1, 2, or more than two studies in 30.1%, 17.4%, and 14.2% of cases, respectively. Patients in Group 2b comparatively underwent 1, 2, or more than two additional studies in 37.6%, 9.2%, and 7.8%, respectively (Figure 9).

**Figure 8.**
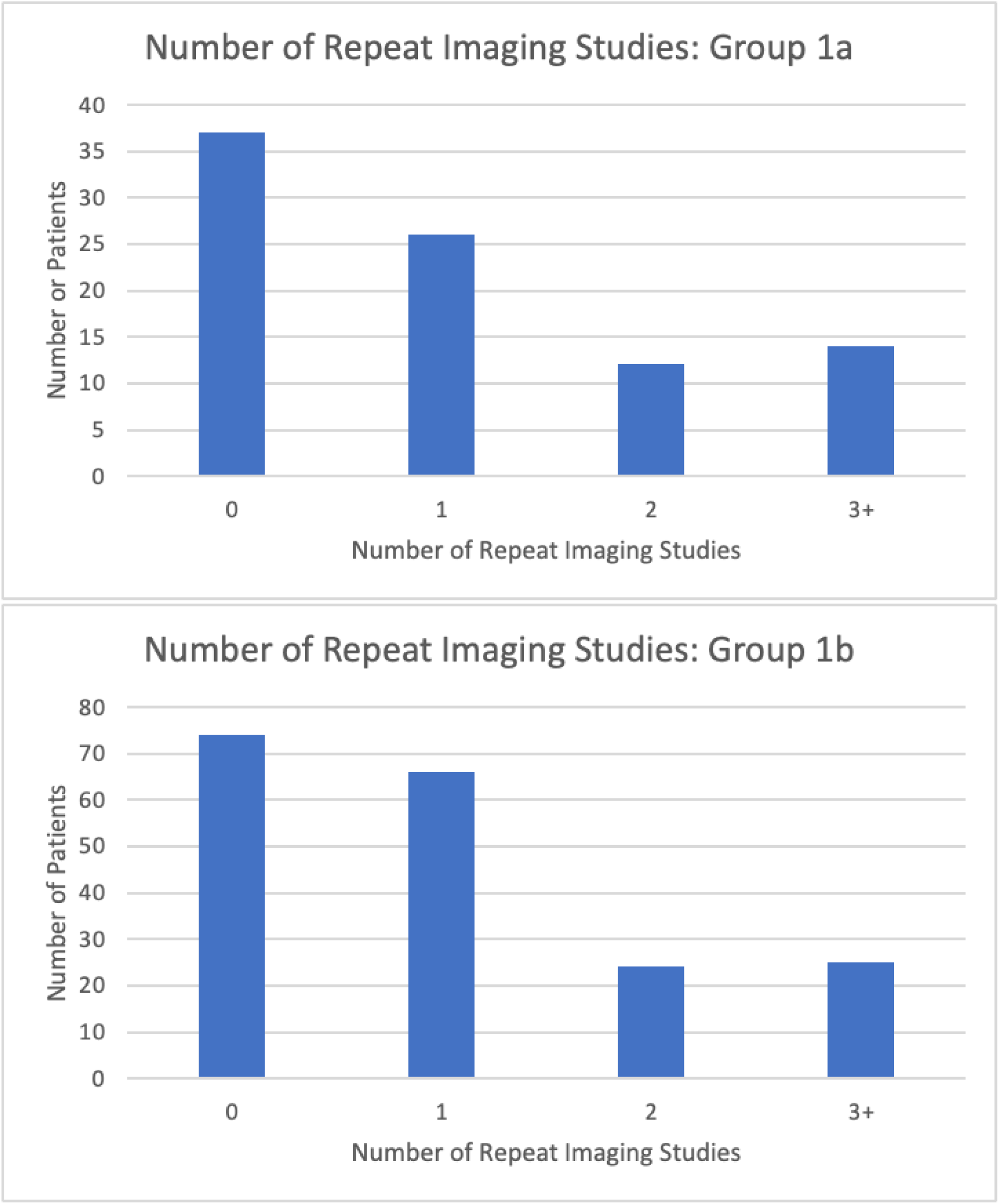
Number of repeat imaging studies for patients in Group 1. The largest group of patients did not obtain any follow-up imaging studies, but 58.4% of patients in Group 1a (A) and 60.9% of patients in Group 1b (B) had at least one follow-up imaging study taken. The average length of time between repeat imaging studies was 100.7 days for patients in Group 1a (A) and 69.3 days for patients in Group 1b (B).

**Figure 9.**
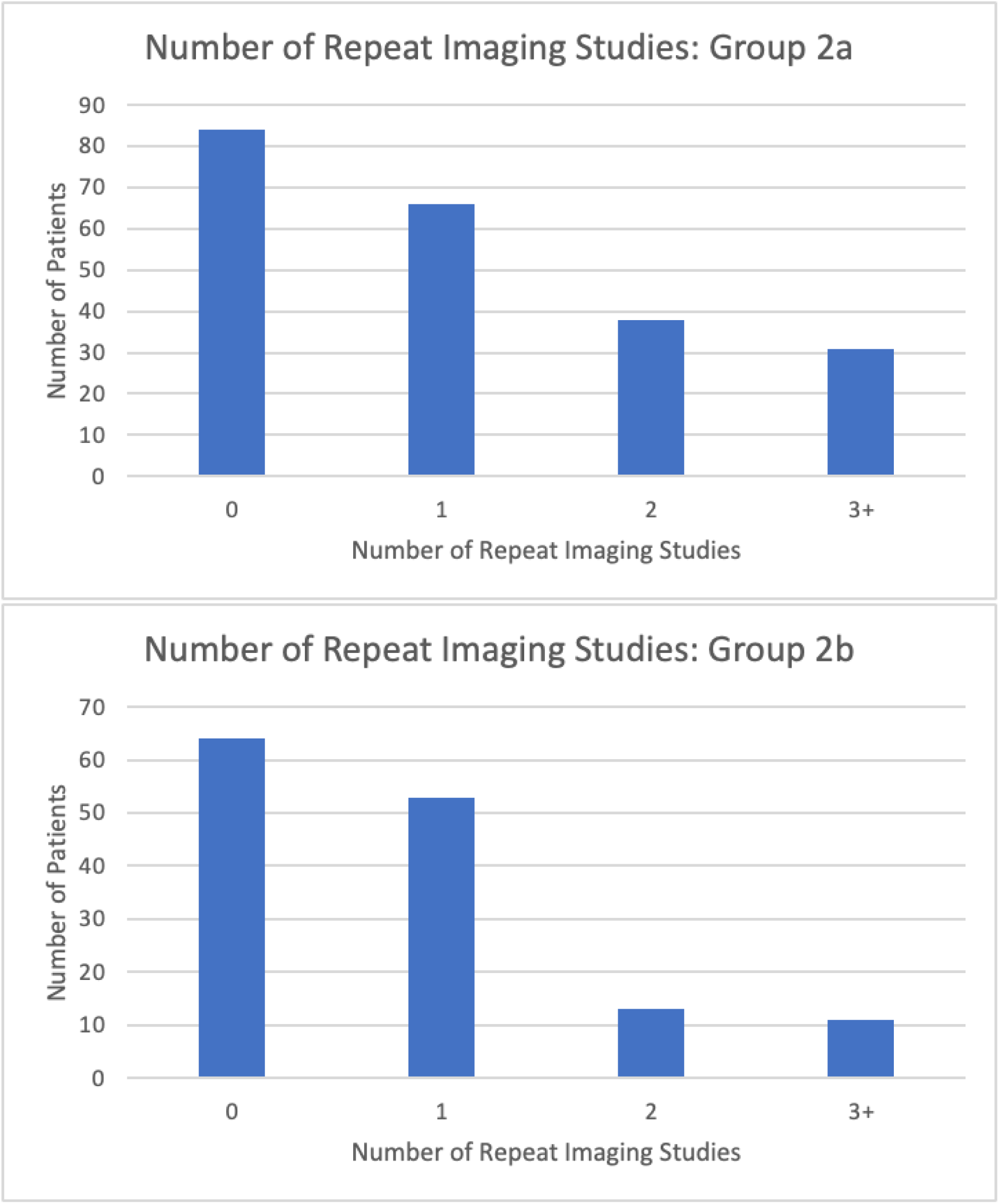
Number of repeat imaging studies for patients in Group 2. 61.6% of patients in Group 2a (A) and 54.6% of patients in Group 2b (B) had at least one follow-up imaging study taken. The average length of time between repeat imaging studies was 90.3 days for patients in Group 2a (A) and 47.3 days for patients in Group 2b (B).

## Discussion

### Background and rationale

Incidental benign bone lesions represent a common diagnostic dilemma for orthopedic oncologists due to their unclear clinical nature, the anxiety they provoke, or the medical-legal concerns that a practitioner may harbor [3, 9, 26]. As a result, they often command resources, time, and money to ensure that the tumor is stable. For example, in a study that surveyed primary care providers (PCPs) managing incidental bone lesions, researchers found that higher patient anxiety levels led some PCPs to pursue further imaging [29]. Patients or family members are not the sole drivers of this practice pattern. Defensive medicine, defined as the use of tests and procedures to avoid or mitigate legal action, has been identified as one of the leading contributors of clinical waste [20]. In a 2012 survey of 72 members of the Pennsylvania Orthopaedic Society, 81% of orthopedic surgeons reported performing defensive imaging over the course of a single day, with nearly 35% of imaging costs attributed to defensive imaging [20]. While there is little hard evidence to prove that this practice is ubiquitous, it is reasonable to suspect that it exists to some degree across the larger health care system. Toward this end, it would be helpful to understand how often benign bone lesions actually progress. Perhaps quantifying the risk, or lack thereof, would alleviate some anxiety for patients. As the lion’s share of benign bone lesions are identified incidentally, it would also be helpful to better understand how often they progress, whether regimented follow-up is warranted, and in turn, what comprises the best practice management strategy.

One approach to limiting practice variation and deviation from optimal utilization of resources is the use of clinical practice guidelines (CPGs), which have been shown to improve clinical decision-making quality as well as patient care consistency [21]. CPGs have become commonplace in numerous disciplines, including medical oncology, radiation oncology, primary care, and pain management, to name a few, and they are often regarded as the gold standard for determining and driving best practices [4, 7, 12–15, 19, 21]. They have repeatedly been shown to significantly improve patient care and health outcomes [21]. A systematic review of the effects of clinical practice guidelines on quality of care across various disciplines found 17 studies that outlined significant improvements in the care given, as well as six studies that reported significant improvements in outcomes [19]. In a report by Blackmore et al., in which 450 physicians’ advanced imaging practices were analyzed after adhering to strict imaging guidelines, the authors demonstrated that they were able to significantly reduce unnecessary imaging utilization using an electronic “decision support system” [4]. Guidelines in the setting of benign bone lesions might represent a valuable tool to prevent excessive follow-up and guide when imaging studies are needed. Currently, no such guidelines exist, but this study’s findings may serve as a foundation for future guidelines on the basis that benign bone lesion progression is an exceptionally uncommon event, particularly in the absence of evolving pain, and less frequent follow-up often suffices.

#### 1. How often do incidentally found bone lesions or bone lesions not requiring surgery at presentation progress over time?

The risk of lesion progression has been explored by a few authors to date. In a retrospective review of 55 patients with long-bone enchondromas, Akoh et al. reported that only 3 of the 55 lesions (5.5%) exhibited subsequent growth. Interestingly, they still recommended surveillance every 3-6 months for the first year and then annually for three years despite the fact that it took 21 months of follow-up before the earliest lesion was noted to have grown [2]. Jassim et al. suggested that patients with incidentally found enchondromas under 5cm do not require any further follow-up if they remain asymptomatic [16]. However, this recommendation was made contingent upon a 2-year period or radiographic stability, thereby supporting some degree of prolonged observation. Conversely, a retrospective review conducted by Ahmed et al. found that only 3 of 58 patients with incidentally found enchondromas exhibited lesional growth. They concluded that follow-up imaging is not required at all for incidental, nonaggressive cartilage tumors and is only recommended if the patient becomes symptomatic [1]. Taken broadly, there is agreement among numerous authors that progression is unlikely and of minimal clinical significance in the absence of symptomatology [1, 2, 10, 16, 22, 23]. Still, there is little consensus concerning length of follow-up. This current study further demonstrates that lesion progression is an extremely uncommon event, especially in the adult population, and corroborates the notion that symptomology is useful to guide management. In the current report, rare cases of progression were often accompanied by clinical symptomatology, indicating that silent progression is a very uncommon phenomenon and may be of little clinical relevance.

#### 2. What is the length of follow-up for patients diagnosed with a benign bone lesion and the frequency of follow-up visits within our institution?

Both asymptomatic and symptomatic patients under observation were managed with a similar number of follow-up visits and imaging studies, suggesting many were unnecessary.

Furthermore, while adult patients are significantly less likely than pediatric patients to experience tumor progression, both populations were followed for similar amounts of time. Though it is widely accepted that skeletal maturity is an important factor in assessing the risk for tumor progression, there is a scarcity of literature comparing follow-up intervals and observational management between adult and pediatric patients. Asymptomatic patients of both groups were also followed for a longer overall period of time, perhaps because it is more difficult to define an endpoint for patients without symptoms, whereas symptomatic patients can improve and provide a rationale for discharging them from care. Our conclusions are concordant with other authors who have proposed that it may be more reasonable for asymptomatic adult patients to return only if and when new symptoms develop, particularly when imaging is entirely consistent with a diagnosis of fibrous dysplasia, non-ossifying fibroma, enchondroma [8].

#### 3. What imaging studies are obtained, and how frequently are repeat imaging studies obtained?

There is also little consensus regarding which imaging modalities should be used for observational monitoring. In a prospective 2016 study looking to examine the most appropriate imaging surveillance in 98 patients with benign intramedullary long bone cartilaginous neoplasms, Kumar et al. concluded that patients with incidental cartilaginous tumors undergo CT scans which can better define calcifications. They also suggested that annual MRIs for at least three years thereafter would be more accurate than annual X-ray imaging [22]. In the same year, Deckers et al. retrospectively reviewed 49 cases of asymptomatic enchondromas and atypical cartilaginous tumors that were followed with annual imaging, and they also recommended patients undergo annual MRIs, which permits for more accurate tumor measurement compared to plain radiographs [10]. Conversely, others have deemed plain radiographs to be adequate for assessing lesional changes in most cases [2, 5, 27, 28]. In that vein, Akoh et al. contested that advanced imaging modalities lead to significant unnecessary expenses and instead recommended using less-costly plain radiographs [2]. Wilson et al. further supported this idea. In a retrospective review of 121 patients with benign cartilage tumors, the authors determined that 85% of patients underwent at least one unnecessary advanced imaging study, and 58% underwent two [27]. Most patients in the present study were diagnosed on plain radiographs, and for 9 of the 10 lesions that grew, the change in size was also noticed on plain radiographs. In the current report, MRI was used primarily for preoperative planning or if plain radiographs were deemed inadequate. In the majority of cases, plain radiographs were found sufficient for observation. Again, some standardization to imaging practices would likely prove helpful and mitigate waste, cost, and risk. While CT scans and plain radiographs are faster and easier to undergo than are MRIs, they do expose patients to ionizing radiation. Exposure over a long-term observational period is particularly relevant since most benign bone lesions affect children and adolescents. Numerous epidemiologic studies on childhood exposure to radiation have demonstrated that even modest doses of radiation are correlated to an increased lifetime risk of cancer in children [6, 17]. Younger patients are more radiosensitive and have more years of life left for cancer to manifest [6]. A 2007 review estimated that up to 2% of US cancers can be attributed to radiation from CT studies, emphasizing how small individual risks applied to an increasingly large population can have major public health implications in the future [6]. Mitigating unnecessary radiation of any kind is recognized as being in the patient’s best interest. Moreover, the average cost of a knee X-ray in the United States is approximately $220 [18]. As previously mentioned, the prevalence of benign bone lesions in asymptomatic children was estimated to be 18.9%, which extrapolates to approximately 13.8 million children under 18 years of age. If each individual obtains just one unnecessary radiograph, that cumulative impact is over $3 billion in preventable costs. Such expenses are likely significantly higher when taking into account more expensive CT and MRI imaging modalities.

## Limitations

Several limitations should be recognized, beginning with all of the limitations inherent in a retrospective review conducted at a single institution. Patients generally presented from a single geographic region and employed management practices may not entirely reflect those of other providers or at other institutions. Multiple histologies were grouped together, which may slightly under or overstate the risk of progression. Additionally, there may have been instances where pain was incorrectly attributed to the adjacent bone lesion or conversely where it was not. The follow-up survey data was limited to those who consented as thus may have limited generalizability and may introduce a potential volunteer bias. With the patient-reported nature of the follow-up survey and the study team’s inability to corroborate patients’ responses, it can not be definitively concluded if those patients had lesions changes. Finally, the study extended over a few years, and is unable to comment on very slow or late progression. The telephone survey extended follow-up an average of 5 additional years, although this was limited to a smaller subset of patients that had originally been lost to follow-up and is therefore not representative of the larger follow-up pattern of this patient population.

## Conclusions

This study describes a single institution’s findings suggesting that excessive follow-up and routine repeat imaging for patients with benign bone lesions may not be necessary. Since a multi-institutional, prospective, longitudinal study is unlikely, the current study may, at minimum, provide reassurance that benign bone lesion progression is an exceptionally uncommon event in the short to intermediate-term. Furthermore, symptomatology almost always heralds lesion progression, so it may be entirely reasonable to advise asymptomatic patients to return for follow-up only when new symptoms develop. While firm and fast guidelines are beyond the scope of this report, the current study may serve as a foundation or perhaps an impetus for future guidelines regarding the observation of benign bone lesions. In the absence of other studies, our findings will hopefully provide assurance to both patients and providers that less frequent follow-up visits and fewer imaging studies often suffice, and limiting them is unlikely to result in inferior outcomes.

## Data Availability

All data produced in the present work are contained in the manuscript

---

**Supplemental Table 1:**
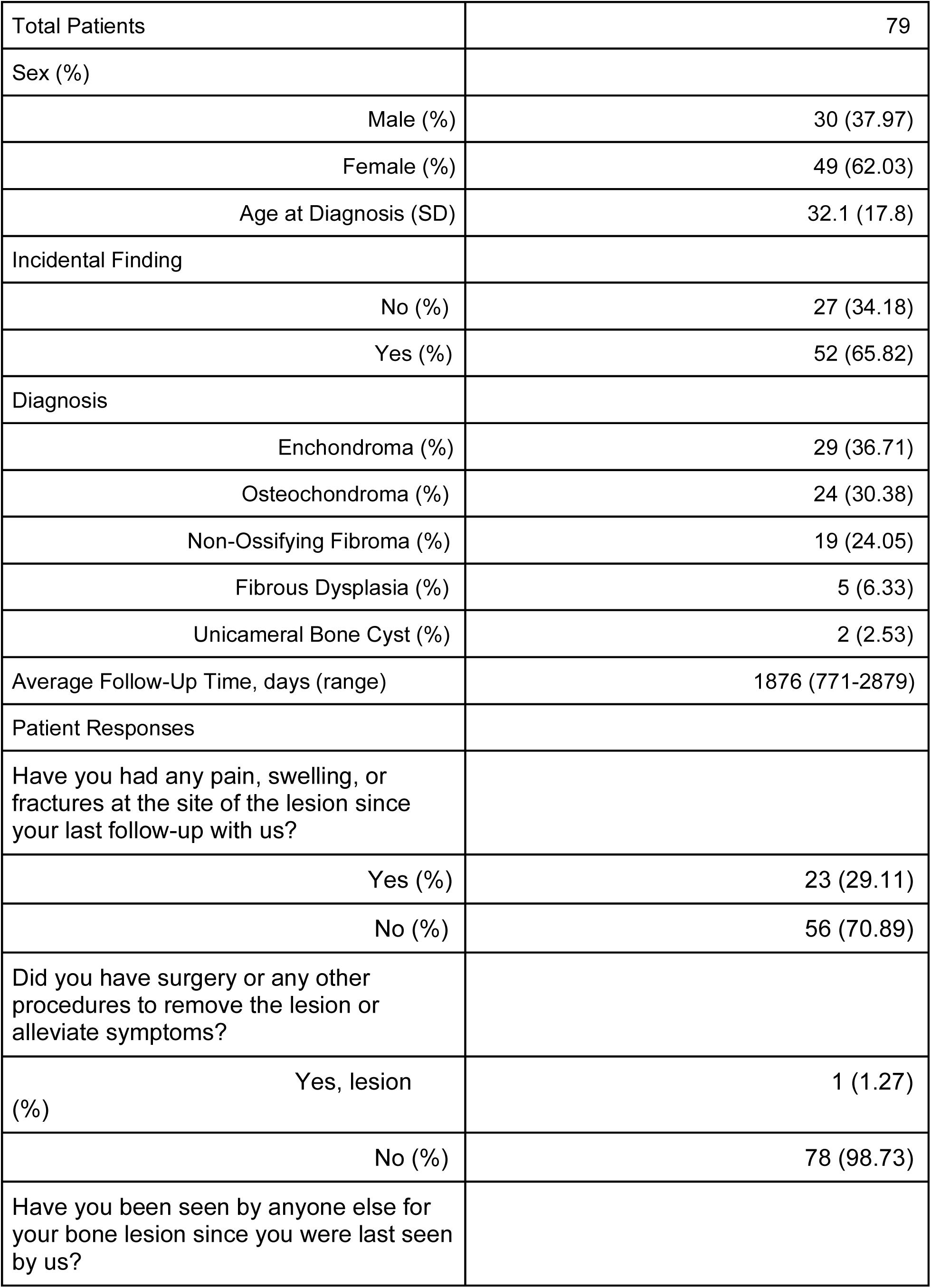
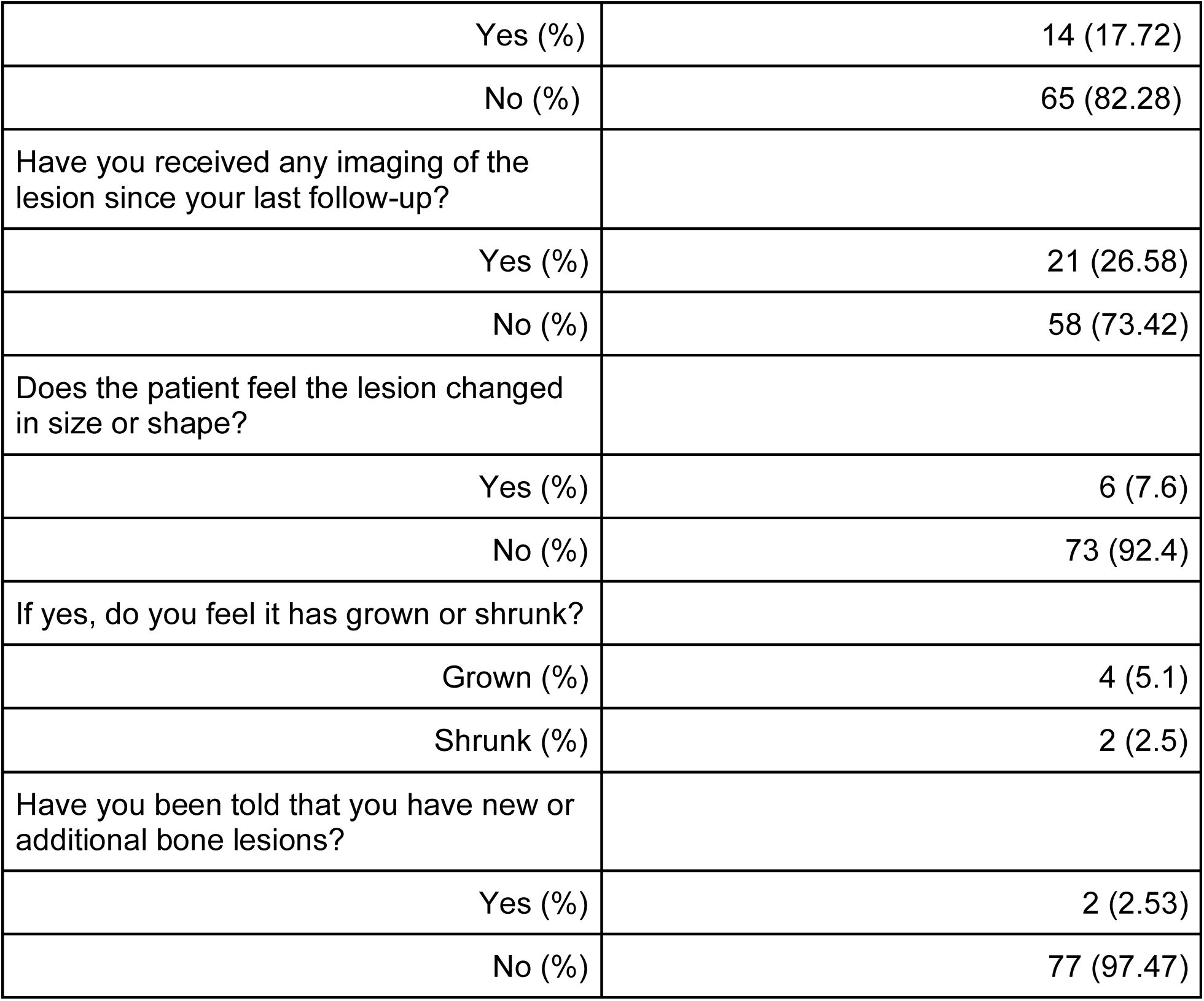

|  |  |
| --- | --- |
| Total Patients | 79 |
| Sex (%) |  |
| Male (%) | 30 (37.97) |
| Female (%) | 49 (62.03) |
| Age at Diagnosis (SD) | 32.1 (17.8) |
| Incidental Finding |  |
| No (%) | 27 (34.18) |
| Yes (%) | 52 (65.82) |
| Diagnosis |  |
| Enchondroma (%) | 29 (36.71) |
| Osteochondroma (%) | 24 (30.38) |
| Non-Ossifying Fibroma (%) | 19 (24.05) |
| Fibrous Dysplasia (%) | 5 (6.33) |
| Unicameral Bone Cyst (%) | 2 (2.53) |
| Average Follow-Up Time, days (range) | 1876 (771-2879) |
| Patient Responses |  |
| Have you had any pain, swelling, or fractures at the site of the lesion since your last follow-up with us? |  |
| Yes (%) | 23 (29.11) |
| No (%) | 56 (70.89) |
| Did you have surgery or any other procedures to remove the lesion or alleviate symptoms? |  |
| Yes, lesion (%) | 1 (1.27) |
| No (%) | 78 (98.73) |
| Have you been seen by anyone else for your bone lesion since you were last seen by us? |  |
| Yes (%) | 14 (17.72) |
| No (%) | 65 (82.28) |
| Have you received any imaging of the lesion since your last follow-up? |  |
| Yes (%) | 21 (26.58) |
| No (%) | 58 (73.42) |
| Does the patient feel the lesion changed in size or shape? |  |
| Yes (%) | 6 (7.6) |
| No (%) | 73 (92.4) |
| If yes, do you feel it has grown or shrunk? |  |
| Grown (%) | 4 (5.1) |
| Shrunk (%) | 2 (2.5) |
| Have you been told that you have new or additional bone lesions? |  |
| Yes (%) | 2 (2.53) |
| No (%) | 77 (97.47) |

